# Impaired Humoral and Cellular Immunity after SARS-CoV2 BNT162b2 (Tozinameran) Prime-Boost Vaccination in Kidney Transplant Recipients

**DOI:** 10.1101/2021.04.06.21254963

**Authors:** Arne Sattler, Eva Schrezenmeier, Ulrike Weber, Alexander Potekhin, Friederike Bachmann, Klemens Budde, Elena Storz, Vanessa Proß, Yasmin Bergmann, Linda Thole, Caroline Tizian, Oliver Hölsken, Andreas Diefenbach, Hubert Schrezenmeier, Bernd Jahrsdörfer, Tomasz Zemojtel, Katharina Jechow, Christian Conrad, Sören Lukassen, Diana Stauch, Nils Lachmann, Mira Choi, Fabian Halleck, Katja Kotsch

## Abstract

Novel mRNA-based vaccines have been proven powerful tools to combat the global pandemic caused by SARS-CoV2 with BNT162b2 efficiently protecting individuals from COVID-19 across a broad age range. Still, it remains largely unknown how renal insufficiency and immunosuppressive medication affect development of vaccine induced immunity. We therefore comprehensively analyzed humoral and cellular responses in kidney transplant recipients after prime-boost vaccination with BNT162b2. As opposed to all healthy vaccinees and the majority of hemodialysis patients, only 4/39 and 1/39 transplanted individuals showed IgA and IgG seroconversion at day 8±1 after booster immunization with minor changes until day 23±5, respectively. Although most transplanted patients mounted spike-specific T helper cell responses, frequencies were significantly reduced compared to controls and dialysis patients, accompanied by a broad impairment in effector cytokine production, memory differentiation and activation-related signatures. Spike-specific CD8^+^ T cell responses were less abundant than their CD4^+^ counterparts in healthy controls and hemodialysis patients and almost undetectable in transplant patients. Signs of alloreactivity promoted by BNT162b2 were not documented within the observation period. In summary, our data strongly suggest revised vaccination approaches in immunosuppressed patients, including individual immune monitoring for protection of this vulnerable group at risk to develop severe COVID-19.

## Introduction

Kidney transplant recipients and patients suffering from kidney failure are imperiled to increased infection risks, either due to dialysis-associated (reviewed in (1)) or therapeutic immunosuppression (IS). This has been comprehensively documented e.g. for cytomegalo-, Epstein-Barr- and BK-virus infection (2), commonly affecting renal transplant recipients with potential implications for allograft function. A growing body of evidence indicates that both patient groups show considerably increased mortality after SARS-CoV2 infection (3-6), arguing in favor of their prioritization in coronavirus disease 2019 (COVID-19) vaccination programs. Large-scale phase III clinical trials (7, 8) have meanwhile demonstrated impressive efficacy of novel mRNA-based vaccines in prevention of severe illness or death. With respect to BNT162b2 vaccine, humoral and cellular responses are documented to be efficiently triggered within one week after boost, with concomitant induction of specific helper and cytotoxic T cell responses (9). Recent data from a BNT162b2 mass vaccination campaign suggest slightly lower effectiveness in patients with comorbidities (10); however, no individual datasets are available for kidney diseases and patients under immunosuppressive therapy were largely excluded from controlled trials. Therefore, accounting for all SARS-CoV2 vaccines authorized thus far, information on kinetics and quality of specific immunity in kidney transplant and hemodialysis patients remains scarce. Experience from influenza A/H1N1 (11, 12) and hepatitis B vaccination trials (13, 14) indicate lower humoral responder rates in both patient groups, likely resulting from combined impairment of early memory B and T cell formation (15). To provide pioneering data on mRNA vaccine-specific adaptive immunity, we quantified humoral and cellular responses induced by BNT162b2 in healthy controls as compared to patients on dialysis and kidney transplant recipients. In the latter group, SARS-CoV2 spike-specific IgG and IgA were rarely detectable, accompanied by broad quantitative and functional impairment of T cell responses. Our study highlights an urgent need to identify alternative or modified immunization strategies for protection of these immunocompromised patients at high risk for SARS-CoV2 associated morbidity and mortality.

## Results

### Study subjects

The study cohort consisted of 39 healthy controls (HC; from which the majority encompassed health care professionals with high vaccination priority), 39 age-matched kidney transplant (KTx) recipients treated with standard immunosuppressive medication, and 26 individuals with kidney failure on hemodialysis (HD). Details of their characteristics are summarized in Table I. Dependent on current vaccination prioritization in Germany, subjects in the HD group exhibited a higher mean age. All individuals were vaccinated with BNT162b2 (Tozinameran) in January or February 2021 with a booster immunization after 21 days. Blood samples for cellular analysis were collected on day 8±1 after boost. Specimen for assessment of humoral immunity were collected for all groups on day 0 and day 8±1 after boost. Sera of 24 KTx patients were additionally analyzed on day 23±5 after boost. None of the study participants had a prior PCR-confirmed SARS-CoV2 infection that was further excluded based on medical history and absence of serum reactivity in a SARS-CoV2 nucleocapsid protein, or, prior to vaccination, spike protein-specific ELISA.

### Absence of vaccination induced humoral immunity in KTx patients

Humoral responses to BNT162b2 vaccination were determined by ELISA. Spike S1 domain-specific IgG reactivity was noted in all 39 healthy controls, 22/26 (84.62 %) dialysis patients, but only in 1/39 (2.6 %) KTx patients at day 8±1 after boost. Comparisons of both patient groups with HC showed significance. Similar findings were made with respect to IgA responses, where solely 4/39 (10.26 %) transplant recipients were sero-reactive as compared to 38/39 (97.44 %) healthy controls and 22/26 (84.62 %) HD patients. Neutralizing antibodies were detected in all 39 healthy individuals, 20/26 (76.92 %) dialysis patients, but in none of the KTx patients examined; comparisons of both patient groups with HC were again highly significant, respectively (Fig. 1A). To decipher whether seroconversion kinetics for transplanted patients were delayed, samples available from 24 previous humoral non-responders were re-analyzed at day 23±5 post booster vaccination. At this timepoint, 2/24 (8.33%) patients showed IgG-and 3/24 (13.04%) IgA seroconversion (Fig. 1B). Relative quantification of spike-specific titers was conducted based on optical density (OD) ratios. Accounting for both isotypes and neutralizing capacity, healthy individuals exhibited significantly higher Ig levels than responding hemodialysis patients (Fig. 1C); due to the low responder rate, statistical analysis for KTx patients was only performed with respect to IgA. Throughout, no signs of acute rejection were observed in KTx patients in response to vaccination during the observation period (d0 to d23±5 after boost), or increased levels of HLA-specific antibodies recorded on day 8±1 after booster immunization as compared to day 0 (Table I).

**Figure 1.**
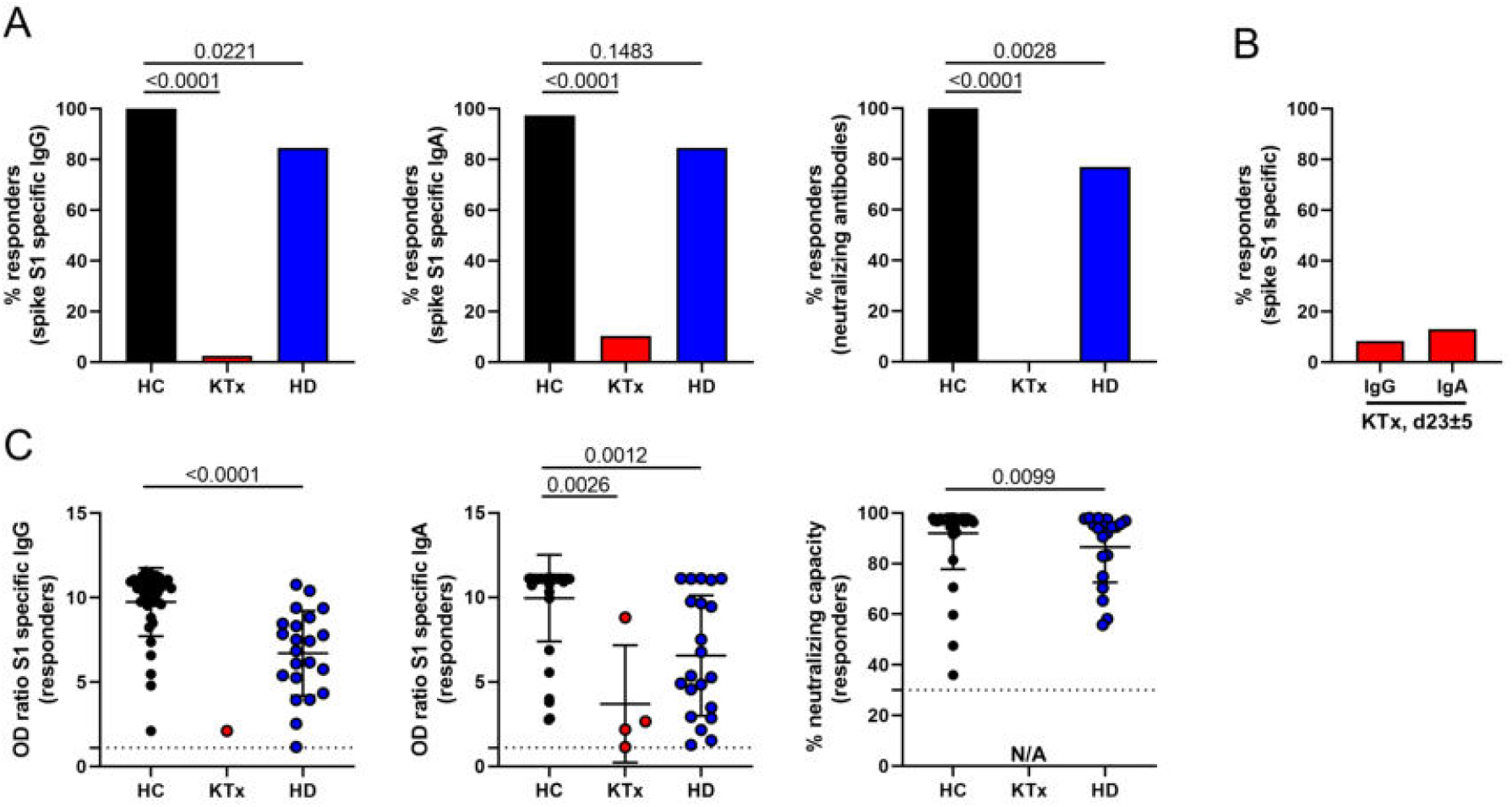
Humoral and cellular reactivity of vaccinees against SARS-CoV2 spike protein. (A) Humoral responder rates were determined based on serum samples collected on day 8±1 after boost being analyzed for spike S1 domain-specific IgG (left, Fisher’s exact test) and IgA (middle, Fisher’s exact test) by ELISA. Surrogate virus neutralization capacity was assessed by a blocking ELISA (right, Fisher’s exact test) with HC: n=39, KTx: n=39 and HD: n=26. (B) Sera of KTx patients available from day 23±5 post boost immunization were re-tested for reactivity as in (A) with n=24. (C) Serological reactivity was quantified only in responding individuals on day 8±1 after boost (IgG (Mann-Whitney-test): HC – n=39, KTx – n=1, HD – n=22; IgA (Kruskal-Wallis-test): HC – n=38, KTx – n=0, HD – n=21; neutralization (Mann-Whitney-test): HC – n=39, KTx – n=0, HD – n=20). N/A – not applicable due to non-responsiveness. Graphs show mean ± SD.

**Table I.** Characteristics of healthy controls (HC), kidney transplant (KTx) and hemodialysis (HD) patients enrolled

| Variable | HC<br>(n=39) | KTx<br>(n=39) | HD<br>(n=26) |
| --- | --- | --- | --- |
| Age (mean yrs ± SD) | 53.03 (17.58) | 57.38 (14.04) | 67.39 (11.88) |
| Females (%) | 19 (48.72) | 11 (28.21) | 9 (34.62) |
| Caucasians (%) | 39 (100) | 39 (100) | 26 (100) |
| **Clinical Parameters** |  |  |  |
| Time on dialysis (mean yrs±SD) | | | 6.87 (5.07) |
| Time since Tx (mean yrs±SD) | | 8.15 (6.09) | |
| Retransplantation (%) |  | 6 (15.38) |  |
| Acute graft rejection (%)* |  | 0 (0) |  |
| HLA antibody induction (%)* |  | 0 (0) |  |
| IS medication |  |  |  |
| CS+Tac+MMF (%) |  | 22 (56.41) |  |
| CS+CyA+MMF (%) |  | 13 (33.33) |  |
| mTORi+MMF±CS (%) | | 3 (7.69) | |
| mTORi+CyA+MMF (%) |  | 1 (2.56) |  |
| Comorbidities |  |  |  |
| Hypertension (%) |  | 37 (94.87) | 22 (84.61) |
| Coronary heart disease (%) |  | 11 (28.21) | 15 (57.70) |
| History of myocardial infarction (%) |  | 4 (10.26) | 4 (15.38) |
| Diabetes (%) |  | 12 (30.77) | 12 (46.15) |
| History of liver disease (%) |  | 4 (10.26) | 9 (34.62) |
| COPD (%) |  | 0 (0) | 3 (11.54) |
| History of malignancy (%) |  | 6 (15.38) | 3 (11.54) |

CS – Corticosteroids, Tac – Tacrolimus, CyA – Cyclosporin A, MMF – Mycophenolate Mofetil, mTORi – mammalian target of rapamycin inhibitor. \*During observation period (d0 to d23 ± 5 after boost).

### Prevalence and magnitude of vaccine-specific T cell responses

For detection of SARS-CoV2 spike glycoprotein or CMV/EBV/Influenza (CEF) control antigen-reactive T cells, PBMC were stimulated with overlapping peptide pools, allowing activation of both CD4^+^ and CD8^+^ T cells in an HLA-type independent manner (16). After pre-gating on live CD3^+^dump^-^ lymphocytes, antigen-reactive CD4^+^ Th cells were identified based on co-expression of CD154 and CD137, as demonstrated earlier (17), allowing sensitive detection with low background (Supplemental Fig.1A). A T cell response was considered positive when peptide mix stimulated cultures contained at least twofold higher frequencies of CD154^+^CD137^+^ (for CD4^+^ T cells) or CD137^+^IFNγ^+^ (for CD8^+^ T cells) cells as compared to the unstimulated control with at least twenty events. The overall prevalence of vaccinees displaying spike-specific CD4^+^ T cell responses was similar for healthy controls, kidney transplant and dialysis patients, ranging from 92-100 %, thereby equalling responder rates to CEF stimulation (Fig. 2A). With respect to the magnitude of the response, however, KTx, but not HD patients, exhibited significantly reduced frequencies of spike-specific CD154^+^CD137^+^ Th cells as compared to healthy controls. This observation did not apply to frequencies of CEF-specific Th cells in transplant recipients (Fig. 2B). Of note, the few transplanted individuals mounting IgA and/or IgG responses until day 23±5 after boost were characterized by significantly higher frequencies of vaccine-specific Th cells than seronegative patients (Fig. 2C).

**Figure 2.**
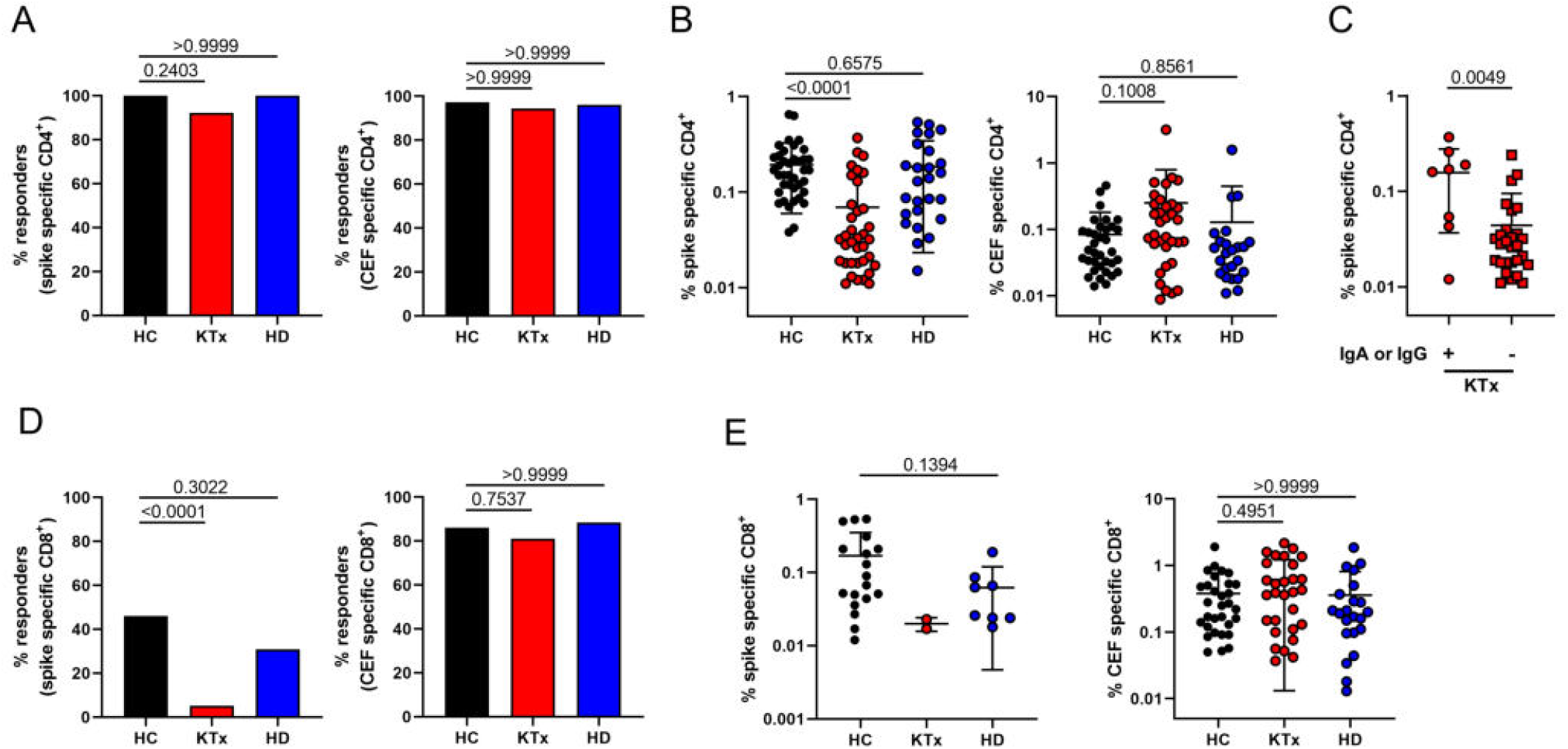
Quantitative features of spike-reactive T cells. (A) PBMC were stimulated with spike (left) or CEF (right) peptide mix for 16 h as indicated. Specific CD4^+^ T cells were identified and quantified by FACS based on co-expression of CD154 and CD137. Depicted are percentages of healthy controls (HC, n=39), kidney transplant (KTx, n=39) and hemodialysis (HD, n=26) patients with a positive CD4^+^ T cell response (responders; Fisher’s exact test, respectively). (B) illustrates frequencies of specific Th cells within responders (HC: spike −n=39, CEF: n=35; KTx: spike −n=36, CEF −n=34; HD: spike −n=26, CEF: n=24; Kruskal-Wallis test, respectively). (C) Portions of spike-specific Th cells in KTx patients showing IgA and/or IgG responses ((+); n=8) or not ((-); n=31; Mann-Whitney test) until day 23±5. (D) Antigen-specific CD8^+^ T cells were identified within PBMC based on coexpression of CD137 and IFNγ. Depicted are percentages within healthy controls (HC, n=39), kidney transplant (KTx, n=39) and hemodialysis (HD, n=26) patients with a positive CD8^+^ T cell response (responders) towards spike (left, Fisher’s exact test) or CEF (right, Fisher’s exact test) stimulation. (E) shows frequencies of spike-(left; Mann-Whitney test) or CEF-specific CD8^+^ T cells (right; Kruskal-Wallis test) within responders (HC: spike −n=18, CEF: n=31; KTx: spike −n=2, CEF −n=30; HD: spike −n=8, CEF: n=22;). Graphs show mean ± SD.

BNT162b2-induced CD8^+^ T cells were identified based on activation-dependent coexpression of CD137 and IFNγ^+^ (Supplemental Fig. 1A). Overall, the prevalence of spike-specific CD8 responses was lower than that determined for CD4^+^ T helper cells with less than 50 % responders within healthy controls and HD patients. Interestingly, vaccine-specific CD8^+^ T cells were detectable only in 2/39 (5.13 %) KTx patients (left), whereas no significant differences between groups were observed for CEF-specific CD8^+^ T cells (right) (Fig. 2D). Frequencies of CD8^+^ T cells in responders to spike stimulation did not significantly differ between HC and HD patients; due to the limited number of responding KTx patients, frequencies were not tested for significant differences to HC. Of note, frequencies of CEF-reactive CD8^+^ T cells did not significantly differ between groups (Fig. 2E).

### Functional repertoire of BNT162b2-reactive T helper cells

Unsupervised analysis using t-distributed stochastic neighbor embedding (t-SNE) of concatenated datasets from all responding patients per group pointed to a reduced production of effector cytokines following spike stimulation in KTx patients as compared to HC and HD patients (Fig. 3A). This finding was reproducible after manual gating, revealing significantly diminished portions of IFNγ, TNFα, IL-2 as well as IL-4 secreting cells in transplanted individuals, whereas only portions of IFNγ secreting cells were diminished in HD patients. Interestingly, frequencies of CEF-activated Th cells from KTx patients were significantly reduced only regarding their IL-2 production capacity (Fig. 3B-E). The ability to co-produce more than one cytokine at a time was then investigated for IFNγ, TNFα and IL-2 with IL-4 being excluded since data were not available for all transplanted patients. Kidney transplant recipients harboured significantly lower frequencies of spike-specific IFNγ^+^TNFα^+^IL-2^+^ (triple^+^) polyfunctional Th cells, associated with an enrichment of cells that produced none of the three cytokines. This observation also applied to polyfunctionality of CEF-specific responses. Frequencies of spike or CEF-specific triple^+^ T cells were not significantly reduced in HD patients as compared to healthy donors (Fig. 3F).

**Figure 3.**
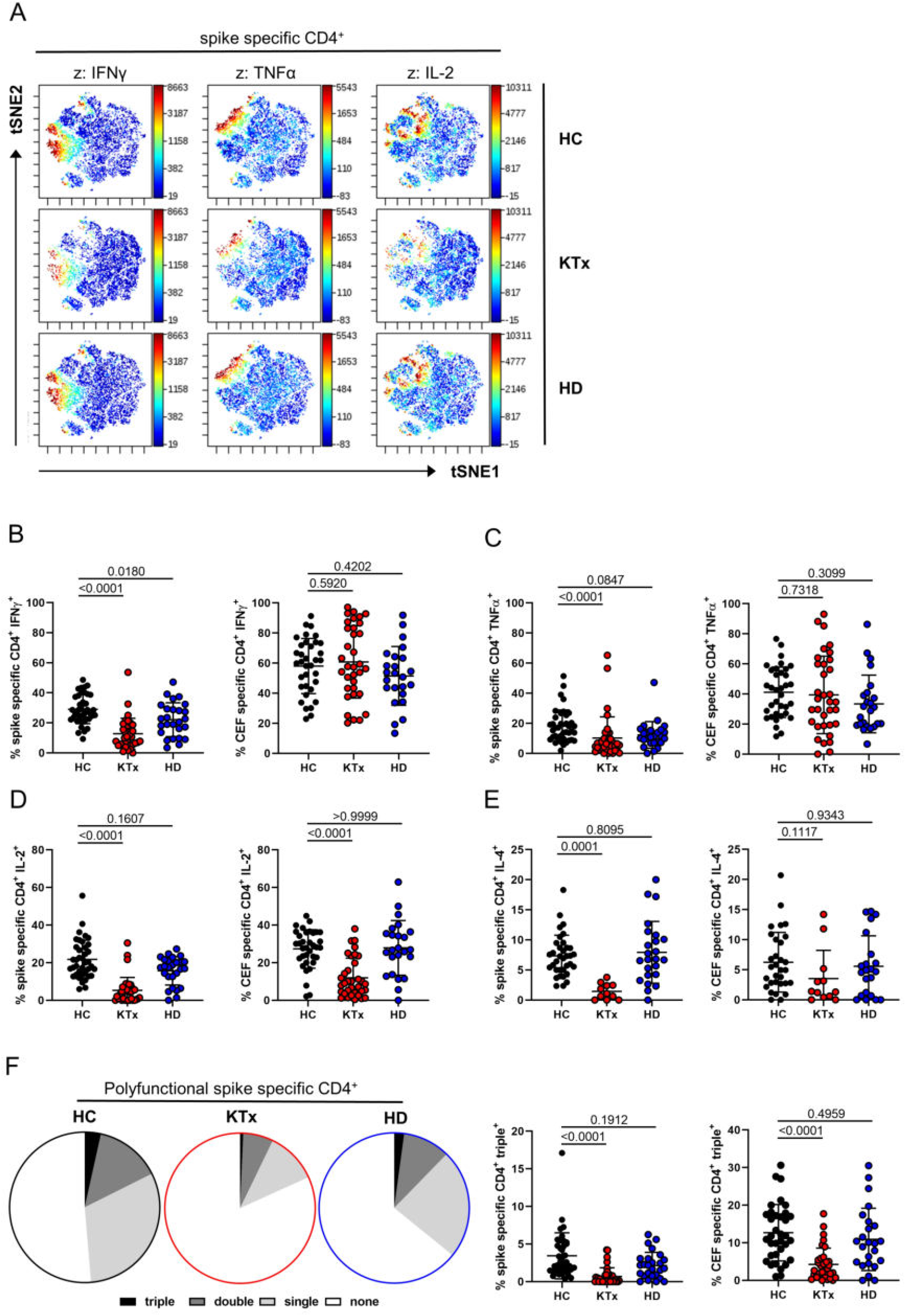
Functional assessment of vaccine-specific CD4^+^ Th cells. (A) Spike-specific CD154^+^CD137^+^ Th cells from all groups were concatenated and subjected to unsupervised analysis using tSNE; highlighted (z-dimension) are areas with IFNγ^+^, TNFα^+^ or IL-2^+^ cells. Spike or CEF-specific CD154^+^CD137^+^ Th cells were further examined after manual gating for expression of (B) IFNγ (spike/CEF: ANOVA), (C) TNFα (spike: Kruskal-Wallis test; CEF: ANOVA), (D) IL-2 (spike/CEF: Kruskal-Wallis test) with n as in Fig. 2B, respectively or (E) IL-4 (spike: ANOVA; CEF: Kruskal-Wallis test; HC: spike −n=35, CEF: n=31; KTx: spike −n=11, CEF −n=12; HD: spike −n=24, CEF: n=22). (F) illustrates (left) the portions of spike-specific T cells expressing three, two, one or no cytokine at a time based on the respective mean values of each group or (right) frequencies of spike or CEF-specific Th cells staining triple positive for IFNγ, TNFα and IL-2 with n as in Fig. 2B and Kruskal-Wallis testing, respectively. IL-4 was excluded from polyfunctionality analyses due to the limited sample size in the KTx group. Graphs show mean ± SD.

### Memory differentiation, ex vivo proliferation and activation state of spike-specific T helper cells

To decipher whether the functional impairment of vaccine-specific Th cells in KTx patients was accompanied by changes in memory formation, subset distribution was analyzed according to expression of CD45RO and CD62L. Whereas the majority of spike-specific Th cells within healthy individuals showed a CD45RO^+^CD62L^-^ effector memory-like (T_EM_) phenotype, their portions were strongly reduced in KTx patients and to a lower, but equally significant extent in HD patients. In both transplant and HD patients, T_EM_ formation impairment was paralleled by a significant increase of short-lived CD45RO^-^CD62L^-^ effector cells. The latter observation also accounted for CEF-specific Th cells in KTx, but not in HD patients (Fig. 4A and B). Overall, spike-specific, as opposed to CEF-specific Th cells showed elevated ex vivo proliferation as reflected by Ki67 expression. Surprisingly, frequencies of Ki67^+^ cells were slightly, but significantly elevated in KTx patients as compared to healthy controls (Fig. 4C). In line with their augmented ex vivo proliferation, spike-but not control antigen-specific Th cells characteristically upregulated the activation/exhaustion associated molecule PD-1 with no marked differences between groups (Fig. 4D). Most spike-specific Th cells expressed the co-activating molecule CD28; in line with data on its downregulation upon frequent encounter with persistent viruses such as CMV (18), transplant recipients harboured slightly, but significantly reduced portions of CD28^+^ CEF-specific Th cells (Fig. 4E).

**Figure 4.**
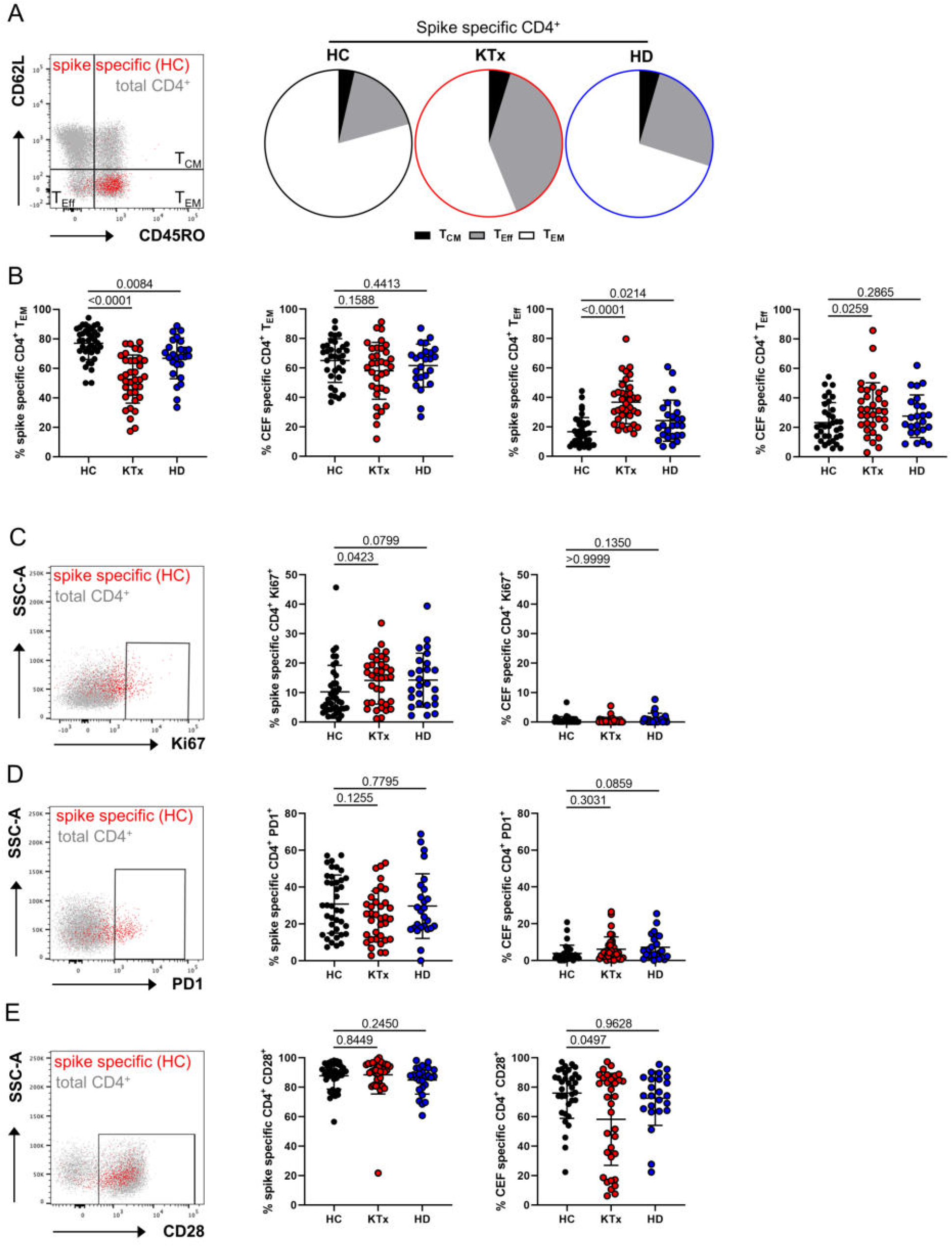
Characteristics of the spike-specific Th cell response with respect to memory formation and ex vivo proliferation/activation. Spike or CEF-specific CD154^+^CD137^+^ Th cells were assessed for their memory or effector phenotype with CD45RO^+^CD62L^-^ identifying effector/memory-type (T_EM_), CD45RO^+^CD62L^+^ central memory (T_CM_) and CD45RO^-^CD62L^-^ effector-like T cells (T_Eff_). (A) Exemplary staining of spike-specific vs. total Th cells from a healthy donor (left) and subset comparison based on the respective mean values for each group (right). (B) summarizes data of spike and CEF-specific T_EM_ (left; spike/CEF: ANOVA) and T_Eff_ (right; spike/CEF: ANOVA) with n as in Fig. 2B. Antigen-specific Th cells were further characterized for (C) ex vivo proliferation based on Ki67 expression (spike/CEF: Kruskal-Wallis test), (D) expression of the activation/exhaustion marker PD1 (spike: ANOVA, CEF: Kruskal-Wallis test) or (E) costimulatory receptor CD28 (spike/CEF: Kruskal-Wallis test) with exemplary overlays of spike-specific vs. total T cells (left, respectively) and summarized data for all groups (right, respectively) with n as in Fig. 2B. Graphs show mean ± SD.

### Transcriptome analysis of vaccine-specific Th cells from KTx patients reveals downregulation of pathways involved in immune activation and cytokine signalling

To collect additional information on differential activation signatures between groups, vaccine-specific CD4^+^ T cells from 3-4 individuals per group were sorted to high purity, typically yielding 200 cells (Supplemental Fig. 1B). Low input bulk RNAseq analysis indicated 49 vs.10 highly differentially expressed (absolute log2 fold change >= 1, FDR < 0.05) genes in KTx vs. dialysis patients compared to healthy probands, respectively. Transcripts e.g., for IFNγ, Th1 differentiation-associated IL-12 receptor β2-chain or TRAF3IP2 involved in NFκB signalling were strongly downregulated in transplanted individuals (Fig. 5A). Pathway analysis further revealed overall downregulation of hallmarks associated with cellular activation, including cytokine signalling, inflammatory responses, allograft rejection or glycolysis, whereas TGFβ signalling motifs were upregulated in spike-specific Th cells of transplant patients. Although several gene sets showed similar patterns in dialysis patients as compared to controls, their enrichment scores remained consistently lower (Fig. 5B).

**Figure 5.**
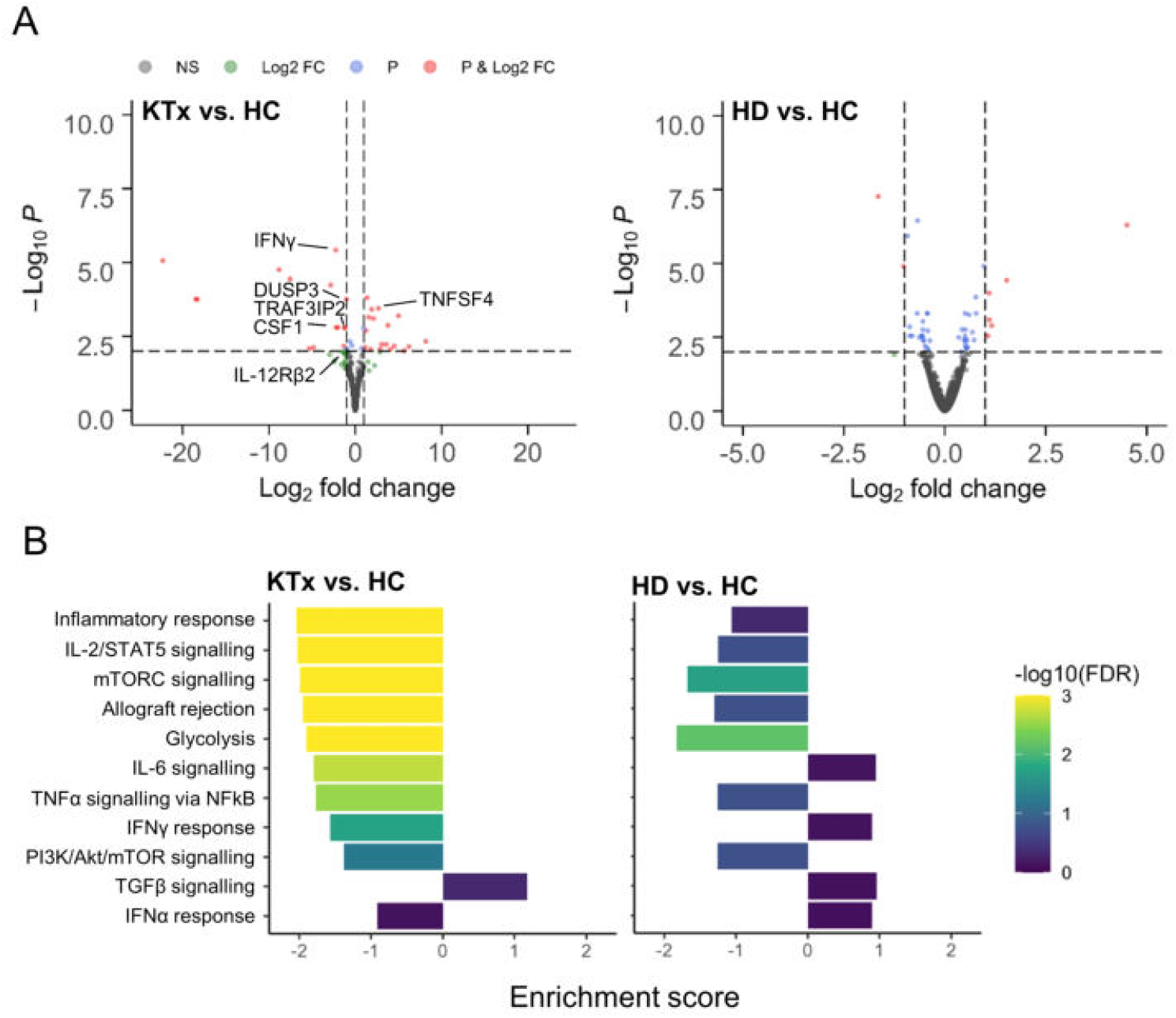
Analysis of differentially expressed genes in vaccine-specific Th cells. (A) Volcano plots depicting the −log10 false discovery rate (FDR) value and log2 fold changes of all expressed genes for comparisons of KTx patients vs. HC (left) and HD patients vs. HC (right). Thresholds for the FDR of 0.01 (“P”) and for the absolute log2 fold change of 1 are indicated by dotted lines, genes passing none (NS – not significant), one or both filters are color-coded. Exemplary genes involved in cellular activation are annotated. (B) Enrichment scores and FDR values for different hallmark gene sets. Direction of the enrichment scores indicates up-or downregulation in the respective comparison. n=3,4,4 for KTx, HD, and healthy individuals, respectively.

### Impact of age and immunosuppressive medication on BNT162b2-induced cellular immunity

Individual predisposition, including age, might strongly impact on anti-viral immunity, as we have recently demonstrated for COVID-19 patients (17). To identify factors that might quantitatively shape vaccine-specific immunity, overall frequencies of CD154^+^CD137^+^ CD4^+^ T cells as well as the ex vivo proliferating Ki67^+^ portion were therefore correlated with age for healthy and transplanted individuals. Frequencies of spike-specific T cells did not correlate with age for healthy controls but showed a trend towards decreased portions with age for KTx patients (P=0.0568). Whereas age in the HC group was positively correlated with frequencies of proliferating Ki67^+^ Th cells, such association was not noted for transplant recipients (Supplemental Fig. 2A). Furthermore, we did not identify associations between time since transplantation and frequencies of spike-specific Th cells or those expressing Ki67 (Supplemental Fig. 2B). Since most KTx patients uniformly received triple immunosuppressive medication and therapy mainly differed based on the type of calcineurin inhibitors (CNI), subgroup analysis was performed for individuals receiving Tacrolimus or Cyclosporine A. Throughout, overall frequencies, portions of cytokine^+^, proliferating or CD45RO^-^CD62L^-^ effector-type Th cells did not show significant alterations between groups (Supplemental Fig. 2C) with similar findings after stratification for low (≤ 1 g per day) or high (2 g per day) dose mycophenolate mofetil (MMF) therapy (Supplemental Fig. 2D).

## Discussion

Based on large phase III clinical trials (7) and access to health care institution recordings (10), tremendous data sets are available suggesting high efficacy of SARS-CoV2 vaccine BNT162b2 in preventing severe or fatal COVID-19 even in individuals with comorbidities or advanced age. Particularly the latter aspect has fuelled the hope that, as opposed to e.g. varicella or influenza vaccines (reviewed in (19)), individuals with otherwise blunted vaccination outcomes might benefit from mRNA based constructs. In this study, by assessing anti-SARS-CoV2 mRNA vaccine-specific immunity, we identify a broad impairment of humoral and cellular responses in kidney transplant recipients under standard immunosuppressive therapy. Whereas BNT162b2 was shown to efficiently induce spike-specific IgG and virus neutralisation titers by day 8 post boost in healthy individuals (9), being in line with our observations, only few transplant recipients seroconverted until day 8±1 after re-vaccination with minor changes until day 23±5. The fact that dialysis patients more frequently developed spike-specific humoral responses, although at rates still below healthy controls, matches inferior vaccination outcomes reported after hepatitis B (20) or influenza A/H1N1 inoculation (11). Correlates of protection against COVID-19 are still incompletely understood and likely include immune components beyond neutralizing antibodies with large animal models particularly highlighting the contribution of T cells upon viral re-challenge (21). Recent data from individuals with mild COVID-19 suggest a critical role for early induction of IFNγ^+^ T cells, being associated with rapid viral clearance (22). On that background, our findings on broad quantitative and qualitative constraints of spike-specific Th cells in KTx patients raises the question as to what extend mRNA-based vaccination might confer protection in this vulnerable group. Using comprehensive multiparameter analysis, our data further reveal significant limitations of vaccine-specific Th effector functions in these individuals, applying to all cytokines examined and equally affecting polyfunctionality. T cells secreting multiple effector molecules at a time have gained particular attention due to their association with superior viral control in HIV infected subjects (23) that was further verified for influenza infection (24). In context with vaccination, multipotent Th cells have been correlated with vaccine-induced immunity against tuberculosis (25). The presence of virus-reactive, multipotent T cells in convalescent seronegative individuals suggests a comparable role in protection against SARS-CoV2 (26), with possible implications for its absence in KTx patients.

Extending flow cytometric data, low input transcriptome analysis of vaccine-specific T helper cells from transplant recipients highlighted downregulation of pathways involved in e.g. cellular activation, cytokine signalling and metabolism. Not surprisingly, these hallmarks represent footprints of immunosuppressive medication, as was e.g. shown for impaired IL-2-STAT5 signalling post kidney transplantation (27). Interestingly, IL-2 gene activity is also sensitive to TGFβ signalling (28), reflecting one of the features we found upregulated in Th cells from KTx patients. Amongst single genes, *TNFSF4 (OX40L)* showed increased transcript levels in this patient group; of note, OX40L protein upregulation was demonstrated only in Th cells after suboptimal antigenic stimulation (29) as is expected in immunosuppressed individuals. Calcineurin inhibitors are further known to impact central components of T cell activation, such as NFκB (30) and metabolic pathways including glycolysis (31), both of which being mirrored in our pathway analyses.

Unexpectedly, we found quantity and quality of CEF-specific Th cells almost indistinguishable in immunosuppressed patients and healthy controls except for IL-2^+^ and polyfunctional Th cells. Studies comparing recall responses to influenza infection versus vaccination in transplant recipients indicated that natural pathogen encounter entails much higher frequencies of antigen-specific T cells that consistently exhibited a broader functional repertoire (32), possibly resulting from strong innate co-stimulation. At least with respect to CMV and EBV, control antigen-specific responses in our KTx cohort relied on natural, and most likely, recurrent infection episodes, thereby providing a possible explanation for the enhanced cytokine production capacity towards CEF as compared to spike antigen stimulation.

Within vaccine-specific Th cells, KTx patients showed a distinct expansion of short-lived effector Th cells at the expense of the memory population. Impairment or retardation of memory formation might represent a direct effect of calcineurin inhibitors as has been comparably demonstrated for Th1, Th2 and Th17 responses (33). Accounting both for functional repertoire and memory development, we cannot exclude different kinetics of vaccine-specific responses in patients as compared to healthy individuals since few KTx patients mounted humoral responses between day 8 and 23 post boost. Delayed mounting of specific T cell responses has been documented for dialysis patients after HBV vaccination, where both cytokine secretion capacity and memory formation normalized at later time points (20).

Whereas responder rates for CD4^+^ T helper cells were comparable between HC and both patient groups in our study, spike-specific CD8^+^ T cells were only detectable in 2/39 (5.13 %) transplant recipients. Both vaccination and infection models have elegantly highlighted the importance of CD4 help for optimal development of memory CD8^+^ T cell responses (34, 35) with CD4 derived IL-2 secretion being key for optimal CD8 priming and effector molecule synthesis (36). The fact that IL-2 production by spike-specific Th cells in KTx patients was strongly impaired, accompanied by downregulation of IL-2-and other cytokine signalling pathways as suggested by RNASeq, may explain, in concert with direct effects of immunosuppressive therapy, the absence of vaccine-specific CD8^+^ T cells in these individuals.

In summary, we demonstrate here that despite advanced mean age and comorbidities, the majority of dialysis patients mounted humoral and cellular responses differing only in select features from healthy individuals. More importantly, however, our data have important implications for vaccination of immunosuppressed individuals, suggesting larger studies to address how vaccine type, dosage and/or number of re-vaccinations might impact successful mounting of antiviral immunity. Given the unexpectedly poor outcome of mRNA-vaccine induced responses in KTx patients, urgent action appears appropriate, affecting not only transplant recipients, but also individuals with other medical conditions requiring immunosuppressive therapy.

## Methods

### Study subjects and assessment of humoral immunity

Demographics of BNT162b2 (Tozinameran, BioNTech/Pfizer, Mainz, Germany) vaccinated healthy individuals and patients that had no history of PCR confirmed SARS-CoV2 infection are summarized in Table I. Previous SARS-CoV2 infection was further excluded by medical history in combination with a negative SARS-CoV2 nucleoprotein-specific ELISA and/or a negative SARS-CoV2 S1 IgG ELISA pre-vaccination (Euroimmun, Lübeck, Germany). Vaccine-specific humoral immunity was assessed in serum samples by ELISA based analysis of SARS-CoV2 spike S1 domain-specific IgG and IgA (Euroimmun, Lübeck, Germany). Samples were considered positive with OD ratios of ≥1.1 as per manufacturer’s guidelines. For examination of virus neutralization capacity, serum samples were analyzed using a surrogate SARS-CoV2 neutralization test (“sVNT”, GenScript, Piscataway Township, USA). The blocking ELISA based assay qualitatively detects anti-SARS-CoV2 antibodies inhibiting the interaction between receptor binding domain (RBD) of the viral spike glycoprotein and angiotensin-converting enzyme. According to the manufacturer’s protocol, inhibition scores ≥30% were considered positive. HLA antibody screening pre-and post-vaccination was performed on a Luminex platform with LABScreen Mixed and Single Antigen Beads (One Lambda). Values above a mean fluorescence intensity (MFI) of 1000 were considered positive.

### Antigens for cellular assays

Stimulations were performed with an overlapping peptide pool consisting of 15-mers with 11 amino acids overlap encompassing the full sequence of the SARS-CoV-2 (GenBank MN908947.3) spike glycoprotein (“Pepmix”, JPT, Berlin, Germany). A combination of overlapping 15-mer peptide mixes including CMV (“Peptivator pp65”, Miltenyi Biotech, Bergisch Gladbach), EBV (“Peptivator consensus”, Miltenyi Biotech) and influenza H1N1 (“Peptivator matrix protein 1”, and “Peptivator nucleoprotein”, Miltenyi Biotech) served as control and is termed CEF throughout. Antigens were used at a final concentration of 1 μg/ml per peptide.

### Cell isolation and stimulation

Serum was collected and immediately cryopreserved. Peripheral blood mononuclear cells (PBMC) were isolated from heparinized blood by Ficoll-Paque™ density gradient centrifugation and cryopreserved in liquid nitrogen. For antigen-specific T cell analysis, 3-5×10^6^ PMBC per stimulation were thawed and washed twice in pre-warmed RPMI1640 medium (containing 0.3 mg/ml glutamine, 100 U/ml penicillin, 0.1 mg/ml streptomycin, 20% FCS and 25 U/ml Benzonase (Santa Cruz, Dallas, USA)), rested for 2 h in culture medium (RPMI1640 with glutamine, antibiotics and 10 % human AB serum, all Biochrom, Berlin, Germany) and stimulated with SARS-CoV2 spike or CEF peptide mix for 16 h. Brefeldin A (10 μg/ml, Sigma-Aldrich) was added after 2 h, enabling intracellular molecule retention. Due to cell number limitations, CEF stimulation was not conducted for all individuals. The same quantity of DMSO contained in peptide mixes was added to the unstimulated control samples.

### Flow cytometric analysis

For detection of surface molecules, antibodies against CD3 (SK7, Biolegend, Carlsbad, CA, USA), CD4 (SK3, Becton Dickinson, Franklin Lakes, NJ, USA), CD8 (SK1, Ebioscience, San Diego, CA, USA), CD45RO (UCHL1, BioLegend), CD62L (DREG-56, BioLegend), PD-1 (EH12.1, Becton Dickinson) and CD28 (CD28.2, Becton Dickinson) were used. Unwanted cells were excluded via a “dump channel” containing CD14^+^ (M5E2, BioLegend), CD19^+^ (HIB19, BioLegend), and dead cells (fixable live/dead, BioLegend). After stimulation, cells were fixed in FACS Lysing Solution (Becton Dickinson), permeabilized with FACS Perm II Solution (Becton Dickinson) and intracellularly stained with anti-CD154 (24-31, BioLegend), anti-CD137 (4B4-1, BioLegend), anti-CD69 (FN50, BioLegend), anti–TNF-α (MAb11, BioLegend), anti–IFN-γ (4SB3, Ebioscience), anti–IL-2 (MQ1-17H12, BioLegend), anti-Ki67 (B56, Becton Dickinson), and anti-IL-4 (MP4-25D2, BioLegend). Cells were analyzed on a FACS Fortessa X20 (Becton Dickinson) flow cytometer.

### Enrichment of spike-specific CD4^+^ T cells, RNAseq and data analysis

For transcriptome analysis, 10^7^ PBMC were stimulated for 16 h with SARS-CoV2 spike glycoprotein peptide mix in the presence of anti-CD40 (1 μg/ml, HB14, Miltenyi Biotec), enabling CD154 surface retention on antigen-reactive cells (37). Thereafter, specific cells were surface stained with anti-CD154 PE (24-31, BioLegend) and magnetically pre-enriched using anti-PE nanobeads (BioLegend) over MACS LS columns (Miltenyi Biotec). Spike-specific CD3^+^CD4^+^DUMP^-^CD154^+^CD69^+^ cells were further sorted in single cell mode to >95 % purity into reaction buffer containing round shaped PCR tube lids on a FACS Aria Fusion cell sorter (Becton Dickinson) and spun down immediately. RNA extraction and cDNA library preparation were conducted with the SMART-Seq® v4 Ultra® Low Input RNA Kit (Takara, Shiga, Japan). Sequencing was performed at the Berlin Institute of Health (BIH) Core Unit Genomics using an Illumina NextSeq 500 platform with 75-bp paired-ends reads. RNAseq reads were trimmed using cutadapt 1.18, retaining reads at least 50 bp long and with at most 10% N content. Following adapter trimming, alignment to the GRCh38 reference genome obtained from ENSEMBL (38) was performed using STAR 2.7.1a (39) retaining only properly paired, uniquely mapping reads. Count matrices were generated using featureCounts from subread 2.0.1 (40) with annotation version GRCh38.98 obtained from ENSEMBL. Downstream processing was performed using DESeq2 1.22.2 (41) in R 3.5.1. Fold changes were shrunk using the ashr method (42). Annotations were added using biomaRt 2.38.0 (43). Differentially regulated pathways between groups were determined by Gene Set Enrichment Analysis (GSEA 4.1.0, (44)) using the hallmark gene set database (45).

### FACS data analysis

FACS data were analyzed with FlowJo 10 (Becton Dickinson). The gating strategy for analysis of antigen-specific T cells is depicted in Supplementary Figure 1A. A T cell response was considered positive when peptide mix stimulated cultures contained at least twofold higher frequencies of CD154^+^CD137^+^ (for CD4^+^ T cells) or CD137^+^IFNγ^+^ (for CD8^+^ T cells) cells as compared to the unstimulated control with at least twenty events. Co-expression of cytokines was analyzed via Boolean gating. Unsupervised analysis was conducted using t-distributed stochastic neighbor embedding (t-SNE) included in Cytobank (Beckman Coulter, Krefeld, Germany). For that, datasets from spike-specific responders were pre-gated in FlowJo on CD154^+^CD137^+^ CD4^+^ cells, followed by concatenation for each group and import into Cytobank.

### Statistics

Statistical examination and composition of ELISA and FACS data derived graphs were executed using GraphPad Prism 8 (GraphPad, La Jolla, CA, USA). Parameter distribution was assessed using The Kolmogorov-Smirnov test. Depending on normal distribution or not, ANOVA (with Holm-Sidak’s post-hoc) or Kruskal-Wallis test (with Dunn post-hoc) were chosen for multiple comparisons. For two-group comparisons, unpaired t test or Mann-Whitney test were used. The relationship between two variables was examined by simple linear regression analysis. For analysis of contingency tables, Fisher’s exact test was applied. In all tests, a value of p<0.05 was considered significant.

## Study approval

The study protocol was approved by the ethics committee of the Charité-Universitätsmedizin Berlin (EA4/188/20), Universitätsmedizin Greifswald (BB 019/21), and Sachsen-Anhalt (EA7/21) and carried out in compliance with its guidelines. All participants gave written informed consent in accordance with the Declaration of Helsinki.

## Data Availability

The data that support the findings of this study are available from the corresponding authors upon reasonable request.

## Author contributions

A.S. designed the study, performed research, analyzed data and wrote the manuscript. E.S., U.W., A.P., F.B. and K.B. designed the study and analyzed data. E.S, V.P., Y.B., C.T. and O.H. performed research. L.T., S.L., D.S., N.L., H.S., B.J., T.Z., and K.J. performed research and analyzed data. C.C. and A.D. designed the study. M.C., F.H. and K.K. designed the study and wrote the manuscript.

## Acknowledgements

The authors are grateful to Dr. Michael Moesenthin, Dr. Peter Bartsch (both Dialysezentrum Burg), Dr. Ralf Kühn, Dr. Dennis Heutling (both Dialyse Tangermünde), Dr. Petra Pfand-Neumann (MVZ Diaverum Neubrandenburg) and Dr. Jörg-Detlev Lippert (Nierenzentrum Köthen) for patient recruitment. We further thank the Charité Universitätsmedizin Benjamin Franklin Flow Cytometry Core Facility (M. Fernandes and A. Branco) supported by DFG Instrument Grants (INST 335/597-1 FUGG, INST 335/777-1 FUGG).

The study was supported by a grant from the Sonnenfeldstiftung, Berlin, Germany to Arne Sattler and Katja Kotsch and DFG grants (KO 2270/7-1, KO-2270/4-1) to Katja Kotsch. Eva Schrezenmeier is participant in the BIH-Charité Clinician Scientist Program funded by the Charité-Universitätsmedizin Berlin and the Berlin Institute of Health and received further support (BCOVIT, 01KI20161) from the Federal Ministry of Education and Research (BMBF). Oliver Hölsken was supported by the Heidelberg Bioscience International Graduate School MD/PhD program, Heidelberg University, Germany. Hubert Schrezenmeier received funding from the Ministry for Science, Research and Arts of Baden-Württemberg, Germany and the European Commission (HORIZON2020 Project SUPPORT-E, no. 101015756).

