## Supplemental Figures 1-4 for "Impaired Humoral and Cellular Immunity after SARS-CoV2 BNT162b2 (Tozinameran) Prime-Boost Vaccination in Kidney Transplant Recipients"

Supplemental Figure 1

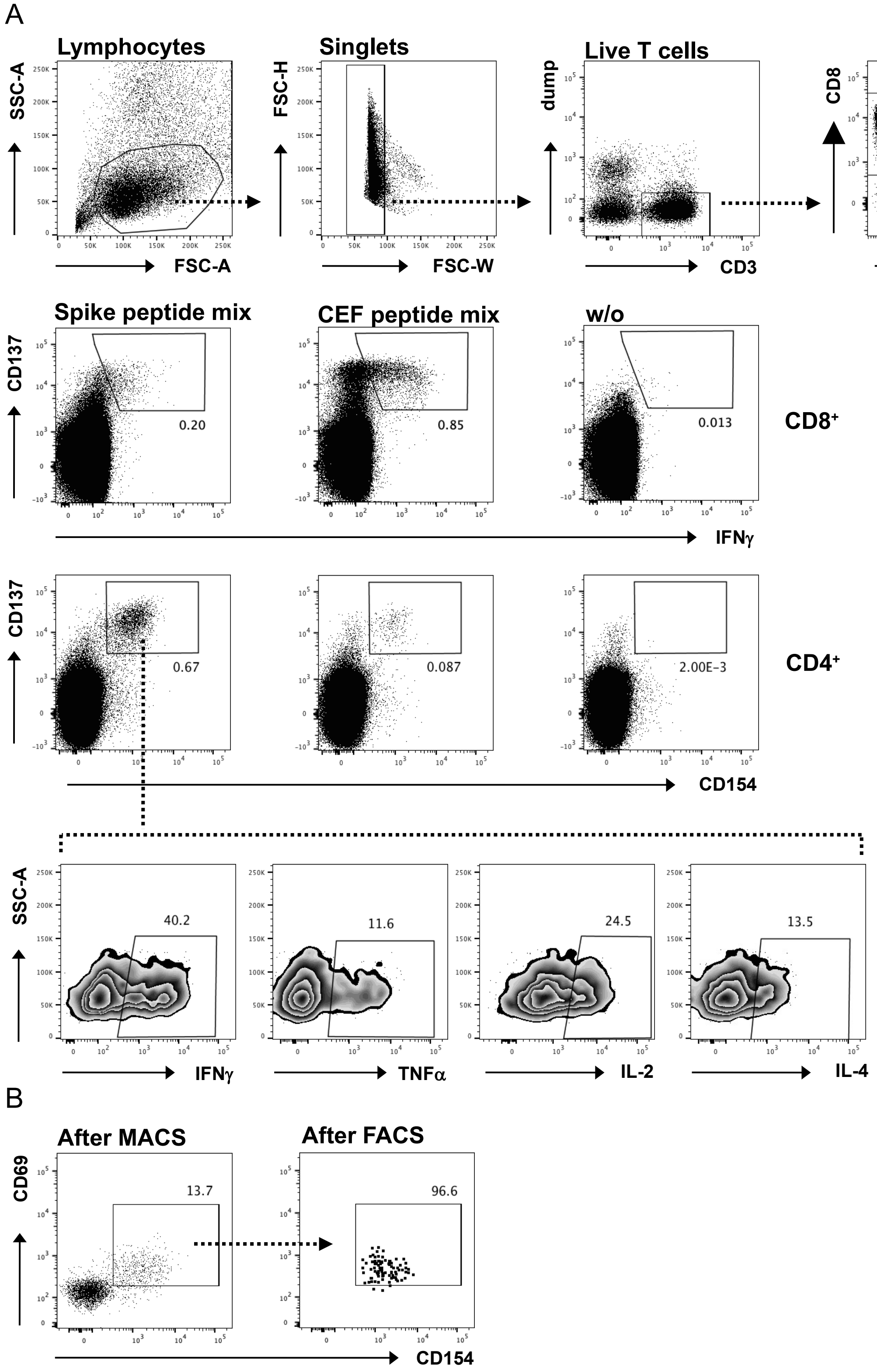


**Identification and enrichment of antigen-specific T cells.** (A) PBMC were stimulated or not with spike or CEF peptide mix for 16 h as indicated. Antigen-specific $CD8^+$ T cells were detected within live single $CD14^-CD19^-CD3^+$ (dump$^-$) lymphocytes according to CD137 and IFNγ co-expression. Live single $CD14^-CD19^-CD3^+$ specific $CD4^+$ Th cells were identified based on co-expression of CD154 and CD137. Specific $CD4^+CD154^+CD137^+$ Th cells were further analyzed for expression of IFNγ, TNFα, IL-2 and/or IL-4. (B) Exemplary reanalysis dot plots depicting pre-enrichment of spike specific $CD4^+$ Th cells via MACS based on activation induced CD154 expression (left), followed by purification via FACS (right).

Impaired Humoral and Cellular Immunity After BNT162b2 Prime-Boost Vaccination in Kidney Transplant Recipients – Sattler et al., 2021

Supplemental Figure 2

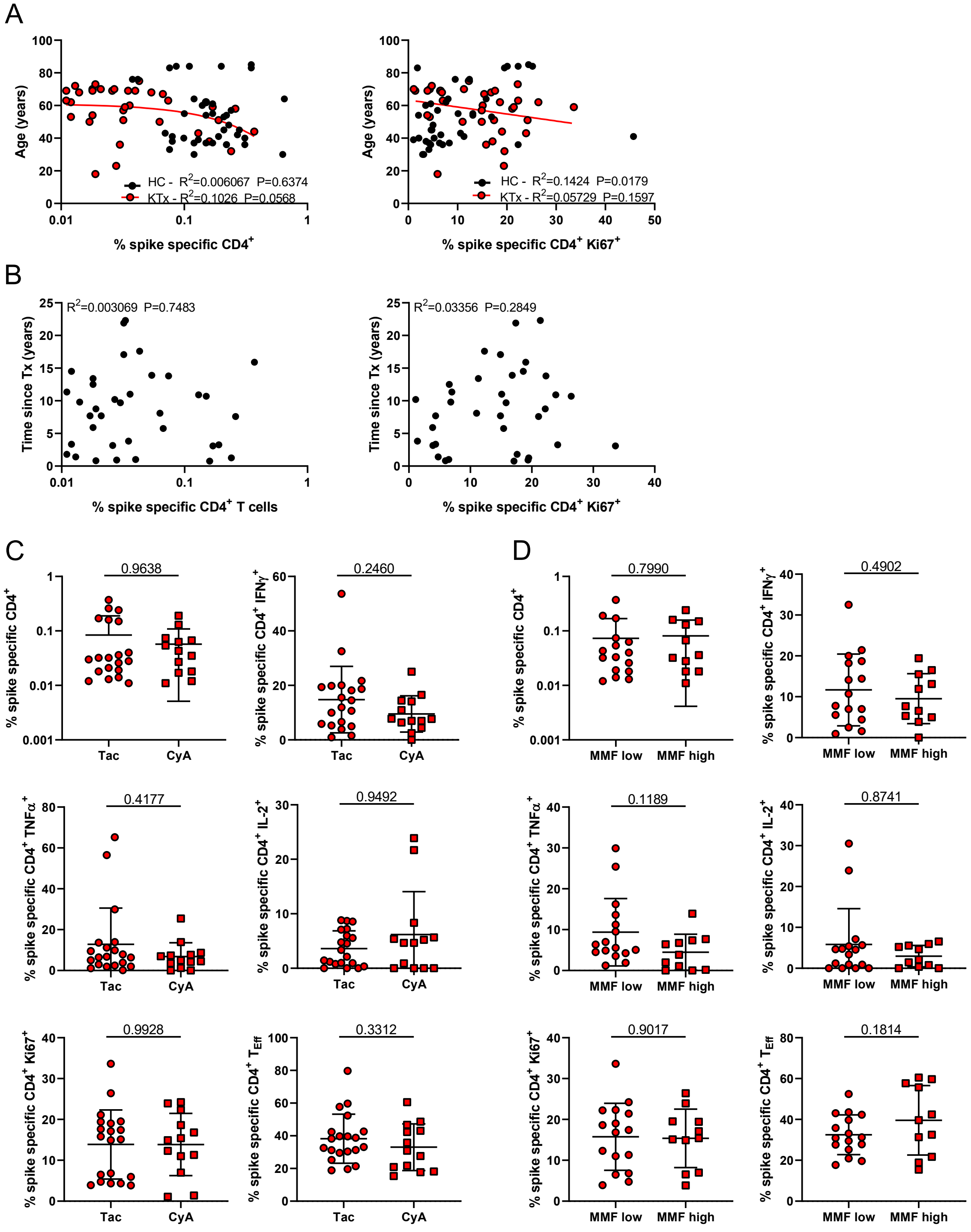


**Association of the vaccine specific $CD4^+$ T cell response with age and IS medication.** (A) Age of HC and KTx patients was correlated with frequencies of spike-specific $CD4^+$ Th cells (left) or frequencies of proliferating $Ki67^+$ cells within the antigen-specific Th cell population (right) with n as in Fig. 2B.
(B) Correlation of time since transplantation with frequencies of spike-specific $CD4^+$ Th cells (left) or frequencies of proliferating $Ki67^+$ cells within the antigen-specific Th cell population (right) in KTx patients with n as in Fig. 2B. (C and D) KTx patients were stratified according to immunosuppressive drug regimen. Frequencies of spike specific Th cells or those expressing IFNγ, TNFα, IL-2, Ki67 or showing a $CD45RO^-$ $CD62L^-$ Teff phenotype were compared in patients receiving Tacrolimus or Cyclosporine A based (C) or low vs. high MMF therapy (D) (Tac: n=20, CyA: n=13, respectively). Graphs show mean ± SD.

Impaired Humoral and Cellular Immunity After BNT162b2 Prime-Boost Vaccination in Kidney Transplant Recipients – Sattler et al., 2021

Supplemental Figure 3

A

| NAME | SIZE | ES | NES | NOM p-val | FDR q-val | FWER p-val | RANK AT MAX | LEADING EDGE |
|---|---|---|---|---|---|---|---|---|
| HALLMARK_UV_RESPONSE_UP | 128 | -0,5641754 | -2,1331058 | 0 | 1,74E-04 | 1,00E-04 | 2415 | tags=37%, list=15%, signal=43% |
| HALLMARK_INFLAMMATORY_RESPONSE | 130 | -0,53510743 | -2,0387063 | 0 | 3,49E-04 | 4,00E-04 | 3152 | tags=42%, list=20%, signal=51% |
| HALLMARK_IL2_STAT5_SIGNALING | 173 | -0,51259583 | -2,0187755 | 0 | 2,33E-04 | 4,00E-04 | 2829 | tags=42%, list=18%, signal=50% |
| HALLMARK_HYPOXIA | 154 | -0,5184372 | -2,0026288 | 0 | 2,13E-04 | 5,00E-04 | 2504 | tags=38%, list=16%, signal=45% |
| HALLMARK_MTORC1_SIGNALING | 193 | -0,49838114 | -1,9945359 | 0 | 2,04E-04 | 6,00E-04 | 2922 | tags=36%, list=18%, signal=44% |
| HALLMARK_ALLOGRAFT_REJECTION | 155 | -0,5003024 | -1,9453664 | 0 | 3,54E-04 | 0,0012 | 2184 | tags=26%, list=14%, signal=30% |
| HALLMARK_GLYCOLYSIS | 155 | -0,49042752 | -1,9036313 | 0 | 5,27E-04 | 0,0021 | 3428 | tags=41%, list=22%, signal=51% |
| HALLMARK_IL6_JAK_STAT3_SIGNALING | 63 | -0,53159034 | -1,7956237 | 2,15E-04 | 0,002151928 | 0,0096 | 1727 | tags=33%, list=11%, signal=37% |
| HALLMARK_KRAS_SIGNALING_DN | 75 | -0,5130576 | -1,7953149 | 6,46E-04 | 0,001929919 | 0,0097 | 1002 | tags=20%, list=6%, signal=21% |
| HALLMARK_TNFA_SIGNALING_VIA_NFKB | 169 | -0,4499193 | -1,770671 | 2,24E-04 | 0,002578845 | 0,0142 | 2525 | tags=33%, list=16%, signal=38% |
| HALLMARK_MYC_TARGETS_V2 | 58 | -0,4802872 | -1,6108648 | 0,005877231 | 0,014407734 | 0,084 | 2769 | tags=34%, list=17%, signal=42% |
| HALLMARK_REACTIVE_OXYGEN_SPECIES_PATHWAY | 46 | -0,49912915 | -1,5893345 | 0,013463627 | 0,016247097 | 0,1021 | 3159 | tags=39%, list=20%, signal=49% |
| HALLMARK_INTERFERON_GAMMA_RESPONSE | 176 | -0,3968431 | -1,5691535 | 6,83E-04 | 0,01840697 | 0,1236 | 3221 | tags=30%, list=20%, signal=37% |
| HALLMARK_ADIPOGENESIS | 165 | -0,39565256 | -1,5510837 | 0,001363017 | 0,02058476 | 0,1473 | 1928 | tags=23%, list=12%, signal=26% |
| HALLMARK_COAGULATION | 59 | -0,46534306 | -1,5489655 | 0,013098561 | 0,019737085 | 0,1509 | 1883 | tags=29%, list=12%, signal=33% |
| HALLMARK_ESTROGEN_RESPONSE_LATE | 126 | -0,40467334 | -1,531774 | 0,002889531 | 0,021827238 | 0,1759 | 2437 | tags=31%, list=15%, signal=36% |
| HALLMARK_COMPLEMENT | 134 | -0,39254677 | -1,4974877 | 0,00447828 | 0,028232267 | 0,2339 | 2885 | tags=30%, list=18%, signal=36% |
| HALLMARK_HEME_METABOLISM | 157 | -0,37600276 | -1,4624645 | 0,005859815 | 0,036552105 | 0,3058 | 2821 | tags=30%, list=18%, signal=36% |
| HALLMARK_CHOLESTEROL_HOMEOSTASIS | 66 | -0,4293268 | -1,4580432 | 0,026105717 | 0,036226396 | 0,317 | 2940 | tags=32%, list=19%, signal=39% |
| HALLMARK_APICAL_JUNCTION | 119 | -0,38424537 | -1,4372096 | 0,010772404 | 0,04122356 | 0,3686 | 2884 | tags=29%, list=18%, signal=36% |
| HALLMARK_MYOGENESIS | 105 | -0,38624853 | -1,421316 | 0,015333333 | 0,045172434 | 0,4122 | 2460 | tags=25%, list=16%, signal=29% |
| HALLMARK_PI3K_AKT_MTOR_SIGNALING | 94 | -0,3814115 | -1,3787745 | 0,034290805 | 0,06129168 | 0,5295 | 3289 | tags=37%, list=21%, signal=47% |
| HALLMARK_ANDROGEN_RESPONSE | 84 | -0,38136986 | -1,3548361 | 0,043639537 | 0,071430475 | 0,6026 | 2829 | tags=26%, list=18%, signal=32% |
| HALLMARK_APOPTOSIS | 135 | -0,34027776 | -1,2972382 | 0,048965055 | 0,10887173 | 0,7705 | 3078 | tags=35%, list=19%, signal=43% |
| HALLMARK_SPERMATOGENESIS | 74 | -0,37096983 | -1,2909025 | 0,08660534 | 0,1097702 | 0,7865 | 3055 | tags=27%, list=19%, signal=33% |
| HALLMARK_MYC_TARGETS_V1 | 200 | -0,30730104 | -1,2327498 | 0,06382979 | 0,1641945 | 0,9099 | 3387 | tags=29%, list=21%, signal=36% |
| HALLMARK_ESTROGEN_RESPONSE_EARLY | 132 | -0,3216158 | -1,2223709 | 0,092887595 | 0,17019427 | 0,927 | 2914 | tags=30%, list=18%, signal=36% |
| HALLMARK_XENOBIOTIC_METABOLISM | 121 | -0,31138954 | -1,1705942 | 0,15408456 | 0,23529126 | 0,976 | 2348 | tags=25%, list=15%, signal=29% |
| HALLMARK_P53_PATHWAY | 169 | -0,28477034 | -1,1138054 | 0,20823902 | 0,32681426 | 0,996 | 1719 | tags=17%, list=11%, signal=18% |
| HALLMARK_KRAS_SIGNALING_UP | 116 | -0,2912415 | -1,0899824 | 0,2715217 | 0,36367878 | 0,9979 | 1949 | tags=20%, list=12%, signal=22% |
| HALLMARK_UNFOLDED_PROTEIN_RESPONSE | 111 | -0,28320986 | -1,0562775 | 0,3199912 | 0,42700946 | 0,9996 | 2618 | tags=22%, list=17%, signal=26% |
| HALLMARK_G2M_CHECKPOINT | 187 | -0,26357728 | -1,0458515 | 0,3312442 | 0,43807837 | 0,9997 | 3374 | tags=27%, list=21%, signal=34% |
| HALLMARK_E2F_TARGETS | 192 | -0,24550505 | -0,977073 | 0,5110102 | 0,60369354 | 1 | 3311 | tags=27%, list=21%, signal=33% |
| HALLMARK_ANGIOGENESIS | 15 | -0,38621876 | -0,9644302 | 0,50950414 | 0,6214266 | 1 | 1921 | tags=27%, list=12%, signal=30% |
| HALLMARK_INTERFERON_ALPHA_RESPONSE | 93 | -0,25187108 | -0,9093883 | 0,6471905 | 0,76174366 | 1 | 397 | tags=4%, list=3%, signal=4% |
| HALLMARK_EPITHELIAL_MESENCHYMAL_TRANSITION | 94 | -0,24942395 | -0,90142405 | 0,67144114 | 0,7628438 | 1 | 2526 | tags=20%, list=16%, signal=24% |
| HALLMARK_PEROXISOME | 81 | -0,25073645 | -0,88513416 | 0,7019462 | 0,78545314 | 1 | 2880 | tags=23%, list=18%, signal=29% |
| HALLMARK_WNT_BETA_CATENIN_SIGNALING | 33 | -0,29142243 | -0,86507404 | 0,6753191 | 0,8135227 | 1 | 4172 | tags=39%, list=26%, signal=53% |
| HALLMARK_NOTCH_SIGNALING | 23 | -0,30740926 | -0,84920996 | 0,68510276 | 0,8279177 | 1 | 1711 | tags=17%, list=11%, signal=19% |
| HALLMARK_APICAL_SURFACE | 23 | -0,3014496 | -0,828369 | 0,7181818 | 0,8480437 | 1 | 4367 | tags=35%, list=28%, signal=48% |

B

| NAME | SIZE | ES | NES | NOM p-val | FDR q-val | FWER p-val | RANK AT MAX | LEADING EDGE |
|---|---|---|---|---|---|---|---|---|
| HALLMARK_UV_RESPONSE_DN | 97 | 0,42865124 | 1,5239751 | 0,00582878 | 0,07493693 | 0,2038 | 2270 | tags=24%, list=14%, signal=28% |
| HALLMARK_BILE_ACID_METABOLISM | 71 | 0,35608506 | 1,2131244 | 0,14803958 | 0,5646358 | 0,9715 | 1132 | tags=15%, list=7%, signal=17% |
| HALLMARK_TGF_BETA_SIGNALING | 47 | 0,3707881 | 1,1713269 | 0,2067803 | 0,50112796 | 0,9919 | 1416 | tags=19%, list=9%, signal=21% |
| HALLMARK_OXIDATIVE_PHOSPHORYLATION | 198 | 0,26288894 | 1,0292147 | 0,3792065 | 0,8755382 | 0,9999 | 2654 | tags=19%, list=17%, signal=23% |
| HALLMARK_FATTY_ACID_METABOLISM | 126 | 0,25521058 | 0,9454126 | 0,5820029 | 1 | 1 | 2510 | tags=20%, list=16%, signal=23% |
| HALLMARK_MITOTIC_SPINDLE | 178 | 0,23568171 | 0,9080746 | 0,7056292 | 0,9987016 | 1 | 3859 | tags=27%, list=24%, signal=35% |
| HALLMARK_PROTEIN_SECRETION | 89 | 0,25184608 | 0,88440883 | 0,70160544 | 0,92864144 | 1 | 2126 | tags=18%, list=13%, signal=21% |
| HALLMARK_DNA_REPAIR | 144 | 0,23132612 | 0,8698067 | 0,7767025 | 0,85021406 | 1 | 2998 | tags=22%, list=19%, signal=27% |
| HALLMARK_HEDGEHOG_SIGNALING | 19 | 0,28498533 | 0,73240435 | 0,83780235 | 0,9576085 | 1 | 2234 | tags=21%, list=14%, signal=24% |

**Gene set enrichment analysis result for genes sorted by differential expression p-value and direction of regulation in KTx vs. HC.** Enrichments for genes (A) down- or (B) upregulated in KTx vs. healthy individuals are shown. ES: enrichment score, NES: normalized enrichment score, NOM p -val: raw p-value, FDR: false discovery rate, FWER: family-wise error rate.

Impaired Humoral and Cellular Immunity After BNT162b2 Prime-Boost Vaccination in Kidney Transplant Recipients – Sattler et al., 2021

Supplemental Figure 4

A

| NAME | SIZE | ES | NES | NOM p-val | FDR q-val | FWER p-val | RANK AT MAX | LEADING EDGE |
|---|---|---|---|---|---|---|---|---|
| HALLMARK_GLYCOLYSIS | 138 | -0,48345116 | -1,8287017 | 0 | 0,007433969 | 0,0054 | 1576 | tags=28%, list=13%, signal=31% |
| HALLMARK_UV_RESPONSE_UP | 115 | -0,47041073 | -1,7274398 | 1,88E-04 | 0,017351985 | 0,0245 | 2675 | tags=45%, list=23%, signal=58% |
| HALLMARK_MTORC1_SIGNALING | 187 | -0,42718133 | -1,6798857 | 0 | 0,020352745 | 0,0433 | 1981 | tags=27%, list=17%, signal=32% |
| HALLMARK_XENOBIOTIC_METABOLISM | 108 | -0,45810518 | -1,6656884 | 1,89E-04 | 0,017777689 | 0,0497 | 1984 | tags=32%, list=17%, signal=39% |
| HALLMARK_MYC_TARGETS_V2 | 57 | -0,49633196 | -1,6382238 | 0,005408538 | 0,019057767 | 0,0663 | 3305 | tags=58%, list=28%, signal=80% |
| HALLMARK_APICAL_SURFACE | 20 | -0,6067829 | -1,6073259 | 0,019530479 | 0,022692421 | 0,0939 | 1573 | tags=30%, list=13%, signal=35% |
| HALLMARK_WNT_BETA_CATENIN_SIGNALING | 28 | -0,55339086 | -1,57638 | 0,017605634 | 0,02668883 | 0,1264 | 2715 | tags=43%, list=23%, signal=56% |
| HALLMARK_MYOGENESIS | 71 | -0,44039714 | -1,5007114 | 0,015748031 | 0,04948565 | 0,2501 | 2166 | tags=35%, list=18%, signal=43% |
| HALLMARK_DNA_REPAIR | 143 | -0,38873306 | -1,4767677 | 0,006228766 | 0,05560669 | 0,3049 | 1621 | tags=22%, list=14%, signal=26% |
| HALLMARK_ESTROGEN_RESPONSE_LATE | 114 | -0,3943081 | -1,4511386 | 0,010638298 | 0,06404827 | 0,3726 | 2018 | tags=29%, list=17%, signal=35% |
| HALLMARK_UNFOLDED_PROTEIN_RESPONSE | 108 | -0,37865734 | -1,3821554 | 0,027573181 | 0,10618592 | 0,574 | 1587 | tags=20%, list=14%, signal=23% |
| HALLMARK_ADIPOGENESIS | 155 | -0,35083774 | -1,3407478 | 0,02631579 | 0,13674612 | 0,6992 | 1896 | tags=27%, list=16%, signal=32% |
| HALLMARK_ALLOGRAFT_REJECTION | 148 | -0,33960602 | -1,29957 | 0,04447795 | 0,1753115 | 0,8118 | 1390 | tags=20%, list=12%, signal=22% |
| HALLMARK_HYPOXIA | 131 | -0,3428638 | -1,2901081 | 0,052820902 | 0,17509621 | 0,8347 | 1968 | tags=27%, list=17%, signal=33% |
| HALLMARK_CHOLESTEROL_HOMEOSTASIS | 62 | -0,38684174 | -1,2877407 | 0,10009681 | 0,16603923 | 0,8396 | 1864 | tags=26%, list=16%, signal=31% |
| HALLMARK_PEROXISOME | 78 | -0,36796567 | -1,2771143 | 0,09290445 | 0,16886579 | 0,8641 | 1229 | tags=17%, list=10%, signal=18% |
| HALLMARK_PI3K_AKT_MTOR_SIGNALING | 91 | -0,356438 | -1,2645499 | 0,09163196 | 0,17515218 | 0,8885 | 2104 | tags=27%, list=18%, signal=33% |
| HALLMARK_TNFA_SIGNALING_VIA_NFKB | 160 | -0,32696941 | -1,2596805 | 0,060977913 | 0,17198606 | 0,8984 | 2895 | tags=38%, list=25%, signal=49% |
| HALLMARK_IL2_STAT5_SIGNALING | 164 | -0,32278168 | -1,2454358 | 0,066753685 | 0,18032515 | 0,9216 | 1478 | tags=19%, list=13%, signal=21% |
| HALLMARK_FATTY_ACID_METABOLISM | 121 | -0,33270532 | -1,2335275 | 0,09382065 | 0,18653947 | 0,9352 | 2493 | tags=26%, list=21%, signal=33% |
| HALLMARK_KRAS_SIGNALING_DN | 55 | -0,35172927 | -1,1464943 | 0,22848894 | 0,32001868 | 0,9943 | 2717 | tags=38%, list=23%, signal=49% |
| HALLMARK_REACTIVE_OXYGEN_SPECIES_PATHWAY | 41 | -0,36580947 | -1,1314687 | 0,26830202 | 0,33564013 | 0,9969 | 2839 | tags=37%, list=24%, signal=48% |
| HALLMARK_INFLAMMATORY_RESPONSE | 121 | -0,28651872 | -1,0637712 | 0,31603774 | 0,4764479 | 0,9999 | 1561 | tags=16%, list=13%, signal=18% |
| HALLMARK_ESTROGEN_RESPONSE_EARLY | 113 | -0,2890921 | -1,0604345 | 0,3254371 | 0,46507168 | 0,9999 | 1992 | tags=27%, list=17%, signal=32% |
| HALLMARK_APICAL_JUNCTION | 93 | -0,28925905 | -1,0374317 | 0,3748053 | 0,5045558 | 0,9999 | 2443 | tags=28%, list=21%, signal=35% |
| HALLMARK_APOPTOSIS | 129 | -0,27663374 | -1,0339795 | 0,37471783 | 0,49388325 | 0,9999 | 2962 | tags=36%, list=25%, signal=48% |
| HALLMARK_HEME_METABOLISM | 143 | -0,26331586 | -0,9976748 | 0,45519096 | 0,5696921 | 1 | 2275 | tags=26%, list=19%, signal=32% |
| HALLMARK_P53_PATHWAY | 161 | -0,2509777 | -0,9701577 | 0,5198954 | 0,6242107 | 1 | 1233 | tags=16%, list=11%, signal=17% |
| HALLMARK_NOTCH_SIGNALING | 19 | -0,35768104 | -0,9342219 | 0,5431927 | 0,70242625 | 1 | 2533 | tags=42%, list=22%, signal=54% |
| HALLMARK_OXIDATIVE_PHOSPHORYLATION | 197 | -0,23607048 | -0,93288046 | 0,64928734 | 0,6828343 | 1 | 1730 | tags=19%, list=15%, signal=22% |
| HALLMARK_COMPLEMENT | 121 | -0,22164218 | -0,8193923 | 0,87114954 | 0,93663484 | 1 | 2094 | tags=22%, list=18%, signal=27% |
| HALLMARK_MYC_TARGETS_V1 | 200 | -0,20483164 | -0,81089294 | 0,9442849 | 0,9229208 | 1 | 1244 | tags=12%, list=11%, signal=13% |
| HALLMARK_SPERMATOGENESIS | 58 | -0,2102307 | -0,6947868 | 0,9622715 | 1 | 1 | 2638 | tags=26%, list=22%, signal=33% |
| HALLMARK_COAGULATION | 46 | -0,21791056 | -0,6867504 | 0,94808745 | 0,97978604 | 1 | 2094 | tags=22%, list=18%, signal=26% |

B

| NAME | SIZE | ES | NES | NOM p-val | FDR q-val | FWER p-val | RANK AT MAX | LEADING EDGE |
|---|---|---|---|---|---|---|---|---|
| HALLMARK_MITOTIC_SPINDLE | 159 | 0,44528213 | 1,737718 | 0 | 0,010422074 | 0,0179 | 1598 | tags=23%, list=14%, signal=27% |
| HALLMARK_UV_RESPONSE_DN | 87 | 0,39546713 | 1,4120328 | 0,029046517 | 0,16643237 | 0,437 | 2316 | tags=32%, list=20%, signal=40% |
| HALLMARK_PROTEIN_SECRETION | 88 | 0,36356315 | 1,3011699 | 0,07064429 | 0,27900046 | 0,7696 | 1707 | tags=24%, list=15%, signal=28% |
| HALLMARK_KRAS_SIGNALING_UP | 99 | 0,34570763 | 1,2596259 | 0,08398727 | 0,28699598 | 0,8692 | 1845 | tags=21%, list=16%, signal=25% |
| HALLMARK_E2F_TARGETS | 182 | 0,30002373 | 1,1899107 | 0,0957782 | 0,3790868 | 0,969 | 2486 | tags=27%, list=21%, signal=34% |
| HALLMARK_G2M_CHECKPOINT | 169 | 0,26614198 | 1,0466211 | 0,3258595 | 0,75955534 | 0,9997 | 2645 | tags=27%, list=23%, signal=35% |
| HALLMARK_ANDROGEN_RESPONSE | 78 | 0,29347807 | 1,0280854 | 0,39508787 | 0,71913296 | 0,9999 | 2284 | tags=28%, list=19%, signal=35% |
| HALLMARK_EPITHELIAL_MESENCHYMAL_TRANSITION | 70 | 0,288305 | 0,98897487 | 0,47003156 | 0,7650182 | 1 | 373 | tags=9%, list=3%, signal=9% |
| HALLMARK_TGF_BETA_SIGNALING | 42 | 0,3073257 | 0,9582165 | 0,521757 | 0,78557426 | 1 | 1187 | tags=17%, list=10%, signal=18% |
| HALLMARK_IL6_JAK_STAT3_SIGNALING | 59 | 0,28399596 | 0,95088637 | 0,53798383 | 0,7305782 | 1 | 1103 | tags=14%, list=9%, signal=15% |
| HALLMARK_INTERFERON_ALPHA_RESPONSE | 92 | 0,25108352 | 0,9030983 | 0,6659677 | 0,8030188 | 1 | 3298 | tags=37%, list=28%, signal=51% |
| HALLMARK_INTERFERON_GAMMA_RESPONSE | 171 | 0,2293831 | 0,9016661 | 0,73079354 | 0,73971325 | 1 | 2977 | tags=32%, list=25%, signal=42% |
| HALLMARK_BILE_ACID_METABOLISM | 61 | 0,24797216 | 0,83061695 | 0,79502714 | 0,83938235 | 1 | 1115 | tags=13%, list=10%, signal=14% |

**Gene set enrichment analysis result for genes sorted by differential expression p-value and direction of regulation in HD vs HC.** Enrichments for genes (A) down- or (B) upregulated in HD vs. healthy individuals are shown. ES: enrichment score, NES: normalized enrichment score, NOM p -val: raw p-value, FDR: false discovery rate, FWER: family-wise error rate.
